# Clinical Characteristics in Children and Adolescents with SARS-CoV-2 Infection: Experience in a highly complex Public Hospital in the city of São Paulo

**DOI:** 10.1101/2020.06.22.20136994

**Authors:** Rafael Santos Rodrigues Vieira, Erisson Linhares de Aguiar, Nara Michelle de Araújo Evangelista, Sergio Antonio Bastos Sarrubbo, Helmar Abreu Rocha Verlangieri, Marcelo Otsuka

## Abstract

**Objective:** Faced with the SARS-CoV-2 epidemic, the real impact of the disease on children and adolescents and the behavior of the disease in this population are questioned. This study aims to assess the clinical characteristics of children and adolescents with SARS-CoV-2 infection and the effectiveness of the measures adopted at the institution.

**Methods:** This is a prospective study carried out from 11/04/2020 to 19/06/2020. Investigated 346 patients between zero and eighteen years old, with analysis of patients diagnosed with COVID-19 confirmed by RT-PCR, obtained from a nasopharynx and oropharynx swab, attended at a highly complex public pediatric hospital in the city of São Paulo.

Protocols for clinical management and treatment of cases of SARS-CoV-2 infection were adopted during the assistance and implementation of a preoperative screening protocol. They were evaluated according to sex, age, epidemiology, presence of comorbidities, clinical manifestations, therapy used, need for hospitalization in the ward and ICU, use of mechanical ventilation (MV) and evolution.

**Results:** 66 confirmed patients with COVID 19 were identified. Median age was 7 years old, with the male gender predominant (2:1). 27 patients (40.9%) had contact with symptomatic respiratory individuals, comorbidity occurred in 50 cases (75.8%). Main clinical manifestations were: fever, 37 patients (56.1%); cough, 23 (34.8%); respiratory distress in 10 (15.2%) and gastrointestinal symptoms in 24 (36.4%). 38 patients (57.6%) received antibiotics and 13 (19.7%) received corticotherapy. 37 patients (56.1%) required hospitalization, eight (19.5%) in the ICU and six (75%) requiring MV. One death occurred and others with good evolution.

**Conclusion:** This study corroborates the perception that the pediatric patient has a more benign manifestation, even in the presence of comorbidities, requiring the screening of surgical patients. The protocol adopted by the institution proved to be effective, with no contamination being observed among patients or between patients and collaborators.

## Introduction

At the end of 2019 a new Coronavirus was identified determining cases of pneumonia in Wuhan, a city in China’s Hubel Province. It spread rapidly, resulting in an epidemic in China, followed by a pandemic affecting up to May 25, 2020, more than 5.5 million people with almost 350,000 deaths worldwide^1^. In February 2020, the World Health Organization designated the disease COVID-19, caused by SARS-CoV-2, due to its similarity to the Coronavirus that determined the severe acute respiratory syndrome in 2002/2003^2^. The virus tends to determine clinical manifestations more frequently in adults and, especially, in the elderly, with high mortality in the population with chronic diseases^3-5^. Most studies confirm the trend towards less severe disease in pediatric patients, and few studies describe the behavior of the virus in children^5-8^.

At the end of February 2020, a public pediatric hospital in the city of São Paulo, to face the announced epidemic, through its clinical staff, was programmed to serve patients with SARS-CoV-2 infection, producing clinical protocols with definition of care flow and therapeutic procedures for patients.

All suspected or confirmed patients with COVID-19, when they need hospitalization, were kept in exclusive sectors destined to the disease, respecting all the criteria for isolation and use of the recommended Personal Protective Equipment (PPE).

## Objectives

In view of the SARS-CoV-2 epidemic, the real impact of the disease on children and adolescents and the disease behavior in this population is questioned, proposing to present the clinical characteristics and the evolution of the disease by SARS-CoV-2 in children and adolescents in a highly complex pediatric service, in addition to evaluating the effectiveness of the proposed measures to deal with the disease in that service.

## Materials and Methods

This is a prospective study carried out from 11/04/2020 to 19/06/2020. A total of 346 patients aged between zero and eighteen years, seen at a highly complex pediatric public hospital in the city of São Paulo, were investigated.

They were investigated for COVID-19 by RT-PCR (obtained from combined nasopharynx and oropharynx swab). Children and adolescents were followed up in specialties or referred from other services for the treatment of other pathologies and always without suspicion of CoViD-19. All were evaluated for sex, age, epidemiology, time to symptom onset and duration, clinical manifestations, need for intensive care, comorbidities and characteristics of the evolution of symptoms and final outcome.

Imaging was obtained whenever there was a clinical sign of pulmonary impairment. Chest tomography (Tc) was performed only in patients who presented any sign considered severe of lung injury (respiratory distress, persistent tachypnea or low O2 saturation without other justifiable causes).

All collaborators who showed any clinical signs or suggestive epidemiology were tested with RT-PCR for CoViD-19, in a total of 163 investigated.

Differentiated flow was determined for patients with epidemiology and / or clinical manifestation suggestive of CoViD-19 infection. The approach of all patients always occurred with the use of recommended PPE. Suspected and confirmed patients, when admitted, were kept in a specific ward for CoViD-19, and assisted by trained teams for care. The ICU was adapted to separate suspected patients in another ward. The entire team was informed about the flows adopted to understand the disease and was advised about protective measures at the institution and outside the institution. A technical reference was created to resolve doubts and all information was made available in the computerized system with free access.

According to guidance from the Pediatric Surgery Society, patients scheduled for essential elective surgeries and urgent surgeries were previously screened with RT-PCR for SARS-CoV-2. Patients undergoing elective surgery were awaiting results at home for effective surgical programming with all the guidelines to avoid contamination. Emergency surgery patients were kept isolated until the result of the RT-PCR.

Descriptive analyzes and a t-test on the number of days of hospitalization were performed to compare patients who had continuous and biphasic evolution and to compare patients who had comorbidities and those who were not exhibited. The level of significance was given by p <0.05. Statistica software was used.

## Results

In the proposed period, 66 patients were identified, with a median age of 7 years (24 days to 18 years old). Male gender was predominant (2:1) and 19.7% (13 cases) below one year of age.

Of the 66 patients, 38 (57.6%) were hospitalized. Of the total confirmed patients, 37 (56.1%) received antibiotics, determined by infection associated with a defined etiology in 7 cases (10.6%), with 4 urinary infections (two Enterobacter cloacae, one E. coli and one K. pneumoniae), 2 appendicitis and a lung culture with Pseudomonas aeruginosa. Antibiotics were also used due to clinical and laboratory criteria without etiological confirmation in 30 patients (45.5%): 20 (30.3%) underwent complementary tests (alteration of leukogram and imaging tests) suggestive of secondary bacterial infection. Ceftriaxone was the most used antibiotic in 17 (25.8%) patients and azithromycin in 12 (18.2%).

Some comorbidity occurred in 50 cases (75.8%), 06 (9.1%) with type 1 diabetes mellitus; 05 (7.6%) with sickle cell anemia; 05 (7.6%) with malformation in the urinary tract; 04 (6.1%) with stage 5 chronic kidney disease; 03 (4.5%) with asthma; 03 (4.5%) with down syndrome; with a history of prematurity, congenital megacolon and patients with epilepsy, each with 02 cases (3%); Five with neoplasms, 02 (3%) with acute myeloid leukemia - M3, 01 (1.49%) with acute myeloid leukemia - M5, 02 (3%) with acute lymphoid leukemia and 01 (1.49%) with tumor of Wilms; marrow aplasia, Leiden factor V deficiency, Pearson syndrome, primary immunodeficiency, nephrotic syndrome, type 3 renal tubular acidosis, Cornelia de Lange syndrome, biliary stenosis, anorectal anomaly and wheezing infant all with 1 case each (1.5 %).

Of the positive patients, 27 (40.9%) had previous contact with confirmed respiratory symptomatic persons, 13 (19.7%) received corticosteroid therapy, three for bronchospasm and one for laryngitis.

Among the patients who required ICU (8), six used mechanical ventilation.

The average length of stay of patients who required an ICU was 36.6 days. In the inpatient unit, 29 patients had some comorbidity and 9 did not have comorbidities, with an average hospital stay of 15.1 and 5.5 days, respectively. There was a significant difference in length of hospital stay between groups with and without comorbidities, with a longer length of stay in the first group. In patients who presented clinical worsening after initial improvement (biphasic evolution), there was a tendency to longer hospital stay and need for ICU.

A death occurred in a patient with severe comorbidity, associated with stage 5 chronic kidney disease, with renal failure and associated infection.

Of the clinical manifestations, fever was present in 37 patients (56.1%), Cough occurred in 23 (34.8%), respiratory distress was in 10 (15.2%) and severe respiratory distress (oxygen saturation in ambient air less than 94%) in 05 children (7.6%) who needed ICU. Gastrointestinal symptoms appeared in 24 patients (35.8%), with vomiting and diarrhea being the most common symptoms presented.

The isolated finding of fever, without other associated symptoms, was identified in 08 (12.1%) patients, and of these, 04 (50%) were infants.

Radiography was obtained in 42 patients with abnormalities in 7 patients. Chest CT was performed on 8 patients with some change in six, with four showing an image suggestive of Coronavirus infection. In two chest Tc findings were compatible with associated bacterial pneumonia.

As of the time of this publication, all patients have evolved with a good clinical improvement of the symptoms presented, except one who has died.

One patient had a clinical suspicion of multisystemic inflammatory syndrome associated with COVID-19. He presented initial symptoms similar to Kawasaki Syndrome (fever for more than five days, alteration of the oral cavity, cervical lymphadenopathy, diffuse maculopapular rash and flaking at the ends of the phalanges). During hospitalization, there was an increase in ferritin, d-dimer and DHL, and treatment for anticoagulation with enoxaparin and the prescription of immunoglobulin and corticosteroids were started. During hospitalization, the patient remained hemodynamically stable, without the need for intensive support, and was discharged for outpatient follow-up.

Characteristics of the population of children and adolescents diagnosed with SARS-CoV-2 infection by sex, age, epidemiology, comorbidities, clinical manifestations, hospitalization, need for ICU and MV, evolution, changes in image and therapy used.

| Characteristics | n | Percentage | p |
| --- | --- | --- | --- |
| Sex |  |  |  |
| Male | 44 | 66,7 |  |
| Female | 22 | 33,3 |  |
| Age |  |  |  |
| < 1 year old | 13 | 19,7 |  |
| Between 1 e 12 years old | 32 | 48,5 |  |
| ≥ 12 years old | 21 | 31,8 |  |
| Epidemiology | 27 | 40,9 |  |
| Clinical manifestations |  |  |  |
| Fever | 37 | 56,1 |  |
| Cough | 23 | 34,8 |  |
| Respiratory distress | 10 | 15,2 |  |
| O2 Saturation < 94% | 8 | 12,1 |  |
| Gastrointestinal manifestation | 24 | 36,4 |  |
| Hospitalization | 38 | 57,6 |  |
| Average days hospitalized | 7,4 |  |  |
| Comorbidities | 50 | 75,8 |  |
| Length of stay (days) |  |  | 0,013 |
| With comorbidities | 9,0 (DP = 14,9) |  |  |
| Without comorbidities | 2,9 (DP = 4,2) |  |  |
| ICU Need | 8 | 12,1 |  |
| MV Need | 6 | 9,1 |  |
| Evolution |  |  |  |
| Continuous | 38 | 57,6 |  |
| Biphasic | 7 | 10,6 |  |
| Asymptomatic | 12 | 18,2 |  |
| Length of stay (days) |  |  | 0,145 |
| Continuous evolution | 8,8 (DP = 15,2) |  |  |
| Biphasic evolution | 9,3 (DP = 5,3) |  |  |
| Asymptomatic | 0,5 (1,2) |  |  |
| Image change | 42 | 63,6 |  |
| Chest radiography | 8 | 12,1 |  |
| Antibiotic use | 37 | 56,1 |  |
| Corticoid use | 13 | 19,7 |  |

The adopted protocol, determining screening for SARS-CoV-2 in all surgical patients, showed 06 (9,1%) positive, all without clinical manifestations.

The analysis of SARS-CoV-2 infection in the clinical staff and patients, did not characterize transmission between patients, and between collaborators and patients. All employees who acquired the infection had important epidemiology outside the institution (163 screened with RT-PCR out of a total of 890 employees with 36 positives, 21 of whom were not workers in the healthcare field).

## Discussion

The SARS-CoV-2 pandemic, different from influenza, but similar to other coronaviruses (SARS and MERS), presents a milder disease in pediatrics, with many asymptomatic or oligosymptomatic pediatric patients^3-7^. However, serious cases, including deaths, also occur in pediatrics and, apparently, the immunological aspect can determine potentially very serious manifestations, such as the Severe Acute Respiratory Syndrome observed in all age groups and others, such as Kawasaki’s Systemic Multisystemic Inflammatory Syndrome^8,9,10^.

So, it is important to understand the behavior of the SARS-CoV-2 infection, its clinical manifestations and evolution in the pediatric patients.

The American Center for Disease Control and Prevention (CDC) mentions that patients affected by COVID-19 can develop mild and moderate symptoms in the vast majority of cases with fever, cough, fatigue, anorexia, dyspnea, sputum production and myalgia. In some situations, presentation with critical manifestations, such as SRAG and involvement of both the myocardium and the Central Nervous System are described^6,7^. In our present study, the manifestations observed in pediatric patients are similar to those observed worldwide, always understanding that these are patients who sought specialized medical service, with associated pathologies in most cases. Most pediatric patients are expected to have mild or asymptomatic manifestations without seeking medical attention. SARS in our study was identified in 10 (15.2%) of the patients, a number considered high compared to other studies in pediatric patients^4,6,8^.

Greater presence in males is also described. In our study, the ratio was 2:1 for males to females.

Almost half of the patients had a history of close contact with a family member who was not adopting quarantine measures, demonstrating that it is still essential to obtain this epidemiological data.

Despite the high percentage that required intervention in the ICU, the patients progressed with good recovery, as shown in publications with pediatric patients^4,6,8^, even in patients with some comorbidity or degree of immunosuppression. In our study, the presence of comorbidity occurred in 75.8% of patients (expected for a highly complex hospital), including cancer patients (6 - AML, ALL and Wilms’ tumor) and with spinal cord aplasia, in addition to patients with chronic use of corticosteroids. Eight patients (12.1%) required an ICU, 75% of whom had mechanical ventilation (9.1% of the total), mainly related to respiratory distress, including the underlying disease (asthma, wheezing infant).

The use of antibiotics (37; 56.1%) was justified by the high risk of infection, as in febrile neutropenic patients, acute chest syndrome or febrile newborns, and / or confirmation or suspicion of infection by clinical and laboratory data. However, etiological isolation occurred in only 07 cases (10.6%).

Adding clinical and laboratory criteria and etiological isolation, concomitant infectious diseases were identified in 37 (56.1%) patients.

Corticosteroid use, despite recent studies demonstrating Dexamethasone as a possibility in the therapy of SARS-CoV-2^11^, occurred in 13 patients (19.7%), but only in one related to the treatment of infection by Coronavirus, by the Multisystemic Inflammatory Syndrome. The others used it due to mechanical ventilation or the underlying disease, such as Asthma and Nephrotic Syndrome.

Among the patients, a 2-year-old child presented a clinical suggestive of Multisystemic Inflammatory Syndrome fulfilling criteria^8,9^, but did not develop shock, as presented in most studies, and with a good response to immunoglobulin and corticosteroids. He presented disease confirmation with positive RT-PCR. Unlike the other reports, we observed a smaller age group, more compatible with classical Kawasaki disease.

Fever occurred in only half of the patients, as described in the literature for pediatric patients, and coughing occurred in about 1/3 of the cases, which are not important clinical manifestations in our study. Fever was not found in critically ill patients, although it was the only symptom among babies.

In most cases, a uniform evolution of the disease was observed, with some cases with biphasic disease (initial improvement followed by worsening). In the seven patients (10.6%) who developed the disease in a biphasic manner, there was a tendency to longer hospital stay (9.3 x 8.2 days), with greater need for ICU and only 06 (10%) patients of those who presented continuous evolution required ICU, with 01 of them evolving to death. This finding brings a reflection of attention in those patients who, after an initial improvement, develop a new clinical worsening.

Patients with some comorbidity tended to have a longer hospital stay (9,0 x 2,9 days), which is expected.

The adoption of RT-PCR screening for SARS-CoV-2 in surgical patients proved to be important, for less risk to the patient, reducing the spread of the disease in the hospital environment, mainly due to the complexity of patients treated at the service (oncological, hematological, illness kidney disease, diabetics, among others), as well as reducing transmission to populations at risk. It is important to highlight the high number of positive patients in the preoperative screening (09 patients).

The determination of SARS-CoV-2 infection in collaborators not directly linked to infected patients, or outside the periods of disease transmission by the patients seen, or in non-care areas, associated with an epidemiology for the disease outside the institution, suggests adequate control with the measures adopted associated with training / continuing education.

## Conclusion

This study corroborates the perception that the pediatric patient has, in most cases, more benign clinical manifestations, even in the presence of comorbidities, but does not rule out the need for strict monitoring, including screening for surgical patients. For our patients who presented severe behavior, the fever was not significant and we emphasize the possibility that the patient who presents an initial improvement may progress later with the possibility of a new worsening, being important the orientation for observing signs of worsening even after discharge from the hospital or outpatient treatment.

The performance of chest CT in no situation changed the conduct or approach adopted. This group considers that the indication for chest CT should be judicious in children and adolescents.

The protocol adopted by the institution demonstrated effectiveness, with no cases of contamination being observed among patients or between patients and collaborators, with a more logical flow and better evidence of acceptance and adoption of measures by the clinical staff, with a reduction in procedural failures, also bringing greater security for the clinical staff. In order to obtain these results, the full participation of the entire clinical team, a multidisciplinary team, in the elaboration of the protocols was essential.

Proposals that aim for better acceptance by the clinical team and mitigate psychological factors, in a moment rarely experienced in the history of medicine, should always be sought, and the sharing of these successful experiences will make this confrontation more effective.

## Data Availability

All data can be obtained in Hospital Infantil Darcy Vargas at Sao Paulo, Brazil.

